# Estimated effectiveness of traveller screening to prevent international spread of 2019 novel coronavirus (2019-nCoV)

**DOI:** 10.1101/2020.01.28.20019224

**Authors:** Katelyn M. Gostic, Ana C. R. Gomez, Riley O. Mummah, Adam J. Kucharski, James O. Lloyd-Smith

## Abstract

Traveller screening is being used to limit further global spread of 2019 novel coronavirus (nCoV) following its recent emergence. Here, we project the impact of different travel screening programs given remaining uncertainty around the values of key nCoV life history and epidemiological parameters. Even under best-case assumptions, we estimate that screening will miss more than half of infected travellers. Breaking down the factors leading to screening successes and failures, we find that most cases missed by screening are fundamentally undetectable, because they have not yet developed symptoms and are unaware they were exposed. These findings emphasize the need for measures to track travellers who become ill after being missed by a travel screening program. We make our model available for interactive use so stakeholders can explore scenarios of interest using the most up-to-date information. We hope these findings contribute to evidence-based policy to combat the spread of nCoV, and to prospective planning to mitigate future emerging pathogens.

## Introduction

As of January 28, 2020, the novel 2019 coronavirus (nCoV) outbreak has been intensifying rapidly in China, and has demonstrated potential for international spread. Many jurisdictions have imposed traveller screening and other travel restrictions (World Health Organization, 2020). It is widely recognized that screening measures are imperfect barriers to spread (Bitar et al., 2009; Gostic et al., 2015; Mabey et al., 2014), due to: the absence of detectable symptoms during the incubation period; variation in the severity and detectability of symptoms once the disease begins to progress; imperfect performance of screening equipment or personnel; or active evasion of screening by travellers. Previously we estimated the effectiveness of traveller screening for a range of pathogens that have emerged in the past, and found that arrival screening would miss 50–75% of infected cases even under optimistic assumptions (Gostic et al., 2015). Yet the quantitative performance of different policies matters for planning interventions and will influence how public health authorities prioritize different measures as the international and domestic context changes. Here we use a mathematical model to analyse the expected performance of different screening measures for nCoV, based on what is currently known about its natural history and epidemiology and on different possible combinations of departure and arrival screening policies.

Our previous analysis considered the contributions of both departure and arrival screening programs, focusing on the context of international spread of infections via air travel. In the current context of the nCoV outbreak, both departure and arrival screening have been proposed and implemented in some countries, though neither approach is likely to be applied uniformly to all air travellers. Traveller screening is also being applied in other contexts, including at roadside spot checks on major routes out of Wuhan. These are directly analogous to departure screens in our earlier analysis, i.e. one-off screening efforts with no delay due to travel duration.

As of January 28, 2020, the Chinese government has been expanding the geographic area and modes of transportation subject to strong travel restrictions. If there was perfect compliance and the restricted area encompassed all areas with community transmission of the virus, then these measures could in theory eliminate the necessity of wider traveller screening. However, multiple factors point to on-going risk, including the existence of substantial numbers of cases in several population centers outside Wuhan (World Health Organization, n.d.), and very early in the outbreak, reports of citizens seeking to elude the restrictions or leaving before restrictions were in place. As the virus continues to spread within China, and as cases continue to appear in other countries, the risk of exportation of cases from beyond the current travel-restricted area is likely to grow.

As a result, increasing emphasis has been placed on the effectiveness of arrival screening to prevent importation of cases to areas without established spread. At the same time, there is great concern about potential public health consequences if nCoV spreads to developing countries that lack health infrastructure and resources to combat it effectively. Limited resources also could mean that some countries cannot implement large-scale arrival screening. In this scenario, departure screening would be the sole barrier -- however leaky -- to importation of cases into these countries. It is also important to recognize that, owing to the lag time in appearance of symptoms in imported cases, any weaknesses in screening would continue to have an effect on case importations for up to two weeks (roughly, the maximum reported incubation period) after changes in screening policy or epidemic context in the source region.

Accordingly, we consider scenarios with departure screening only, arrival screening only, or both departure and arrival screening. The model can also consider the consequences when only a fraction of the traveller population is screened, due either to travel from a location not subject to screening, or due to deliberate evasion of screening.

The central aim of our analysis is to assess the expected effectiveness of screening for nCoV, taking account of current knowledge and uncertainties about the natural history and epidemiology of the virus. We therefore show results using the best estimates currently available, in the hope of informing policy decisions in this fast-changing environment. We also make our model available for public use as a user-friendly online app, so that stakeholders can explore scenarios of particular interest, and results can be updated rapidly as our knowledge of this new viral threat continues to expand.

## Results

### Model

The core model has been described previously (Gostic et al., 2015), but to summarize briefly, it assumes infected travellers can be detained due to the presence of detectable symptoms (fever or cough), or due to self-reporting of exposure risk via questionnaires or interviews. Before screening, travellers can be classified into one of four categories: (1) symptomatic and aware that exposure may have occurred, (2) symptomatic but not aware of exposure risk, (3) aware of exposure risk but without detectable symptoms, and (4) neither symptomatic nor aware of exposure risk (Fig. 1). Travellers in the final category are fundamentally undetectable, and travellers in the third category are only detectable if aware that they have been exposed and willing to self report.

**Fig 1.**
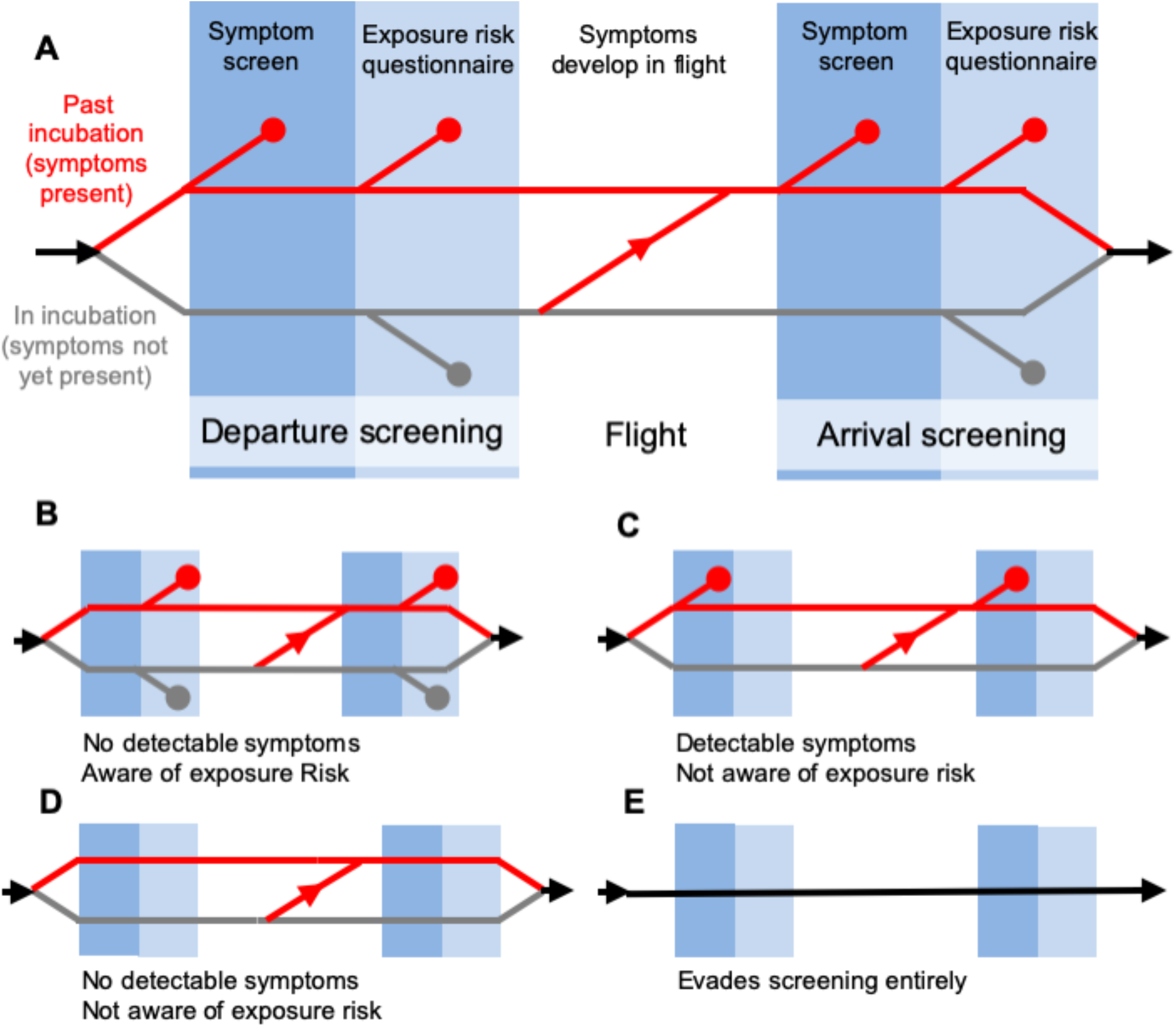
Model of traveller screening process, adapted from Gostic et al., eLife, 2015. Infected travellers fall into one of five categories: (A) Symptomatic cases aware of exposure risk are detectable in both symptom screening and questionnaire-based risk screening. (B) Subclinical and not-yet-symptomatic cases aware of exposure risk are only detectable using risk screening. (C) Symptomatic cases unaware of exposure risk are only detectable in symptom screening. (D-E) Subclinical cases who are unaware of exposure risk, and individuals that evade screening, are fundamentally undetectable.

In the model, screening for symptoms occurs prior to questionnaire-based screening for exposure risk, and detected cases do not progress to the next stage. This approach allows us to track the fraction of cases detected using symptom screening or risk screening at arrival or departure. Additionally, the model keeps track of four ways in which screening can miss infected travellers: (1) due to imperfect sensitivity, symptom screening may fail to detect symptoms in travellers that display symptoms; (2) questionnaires may fail to detect exposure risk in travellers aware they have been exposed, owing to deliberate obfuscation or misunderstanding; (3) screening may fail to detect both symptoms and known exposure risk in travellers who have both and (4) travellers not exhibiting symptoms and with no knowledge of their exposure are fundamentally undetectable. Here, we only consider infected travellers who submit to screening. However, the supplementary app allows users to consider scenarios in which some fraction of infected travellers intentionally evade screening (Fig. 1E).

### Parameters

The probability that an infected traveller is detectable in a fever screen depends on: the incubation period (the time from exposure to onset of detectable symptoms); the proportion of subclinical cases (mild cases that never develop detectable symptoms); the sensitivity of thermal scanners used to detect fever; the fraction of cases aware they have high exposure risk; and the fraction of those cases who would self-report truthfully on a screening questionnaire. Further, the distribution of individual times since exposure affects the probability that any single infected traveller has progressed to the symptomatic stage. In a growing epidemic, the majority of infected cases will have been recently exposed, and will not yet show symptoms. We used methods described previously to estimate the distribution of individual times since exposure for different parameter regimes (Gostic et al., 2015). Briefly, the model assumes the fraction of cases who are recently exposed increases with R0. The distribution of times since exposure is truncated at a maximum value, which corresponds epidemiologically to the maximum time from exposure to patient isolation, after which point we assume cases will not attempt to travel. (Isolation may occur due to hospitalization, or due to confinement at home in response to escalating symptoms or nCoV diagnosis).

At the time of this writing, nCoV-specific estimates are available for most of these parameters, but almost all have been derived from limited or preliminary data sources and remain subject to considerable uncertainty. Table 1 and the Methods summarize the current state of knowledge. Here, we used two distinct approaches to incorporate this uncertainty into our analysis.

**Table 1.**
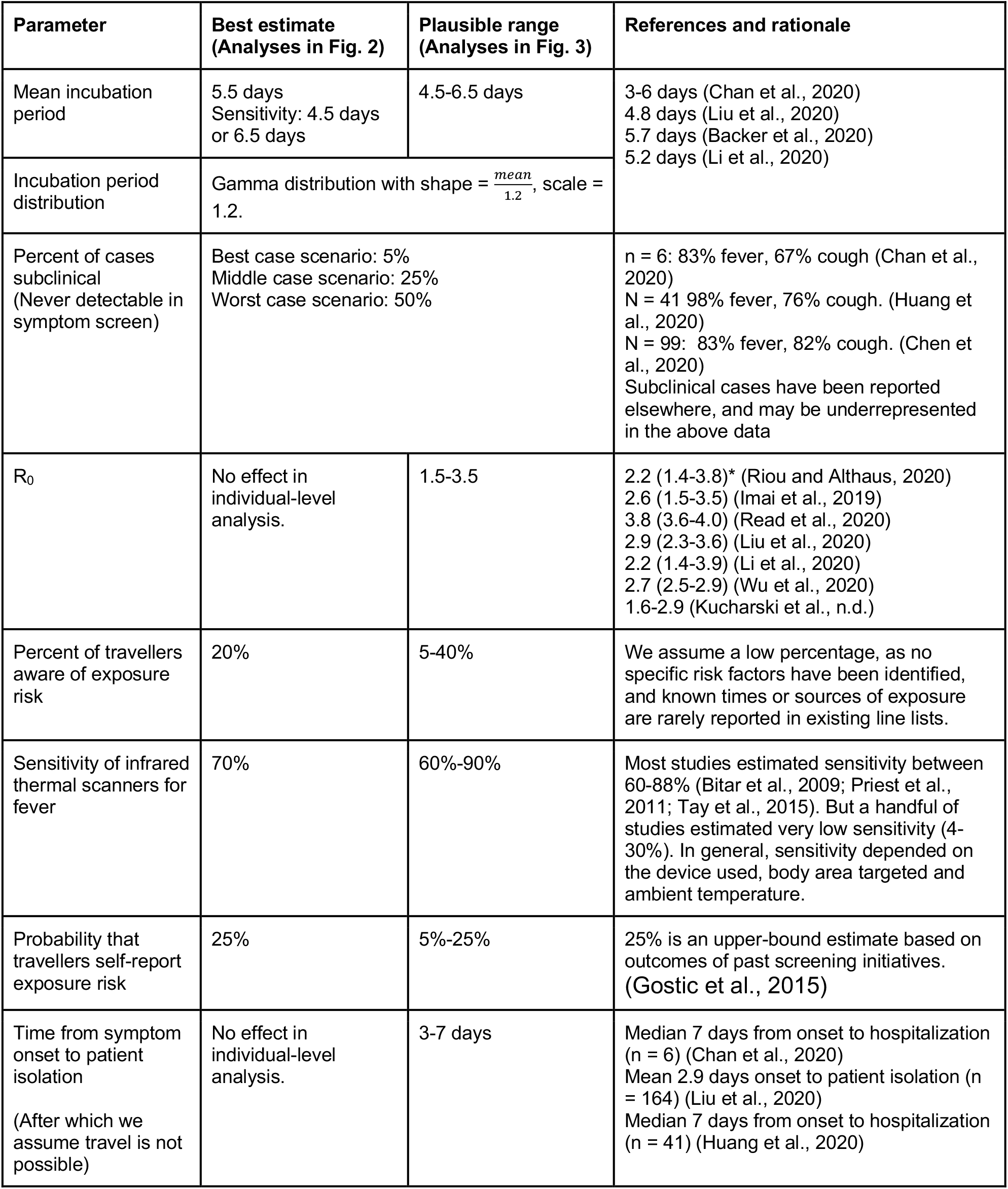
Parameter values estimated in currently available studies, along with accompanying uncertainties and assumptions. *Confidence interval, credible interval or range reported by each study referenced.

First, to estimate the probability that an infected individual would be detected or missed (Fig. 2), we considered a range of plausible values for the mean incubation time, and the fraction of subclinical cases. We focus on these two parameters because screening outcomes are particularly sensitive to their values. All other parameters used to generate Fig. 2 were fixed to the best available estimates listed in Table 1.

**Fig 2.**
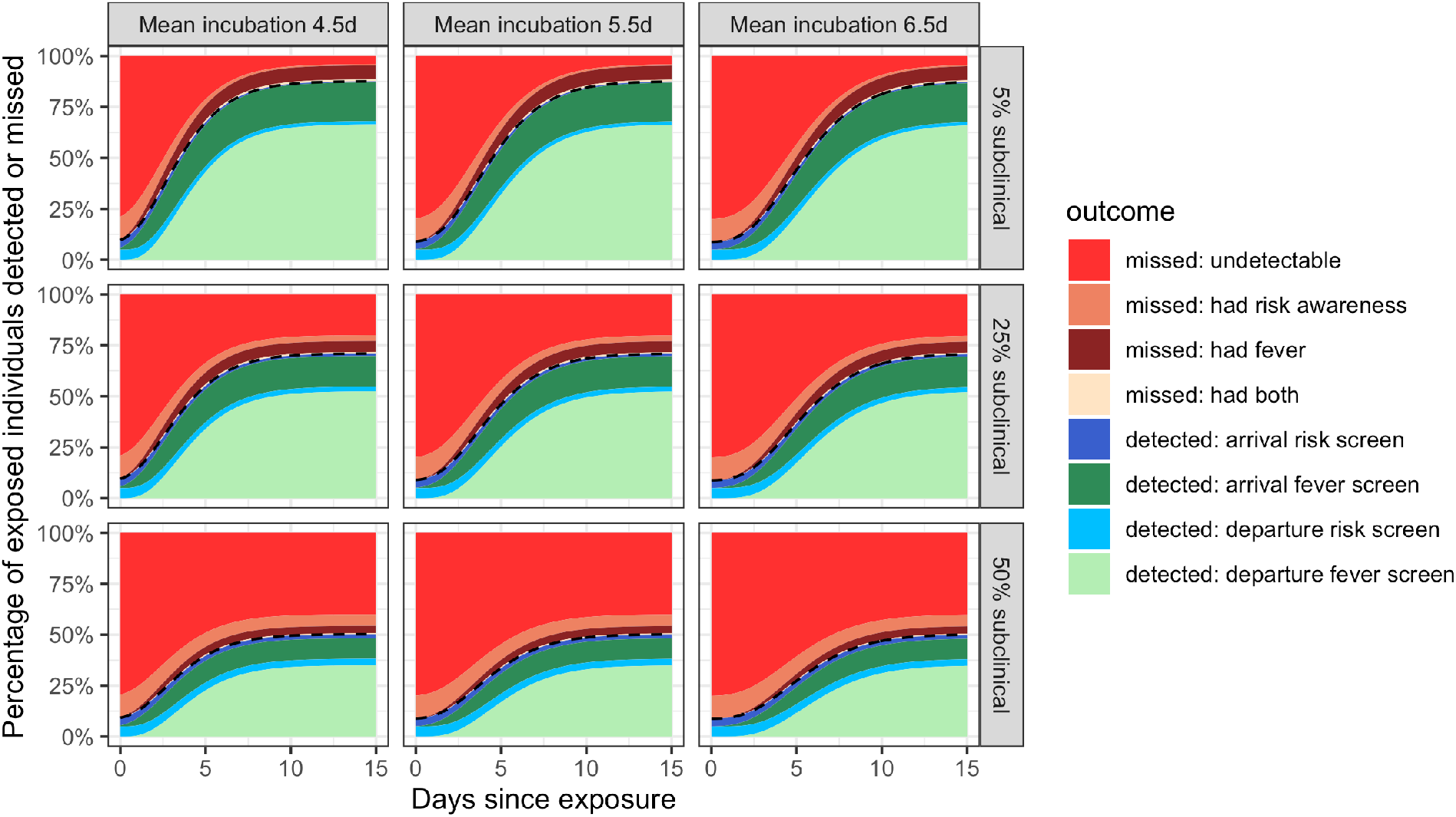
Individual outcome probabilities for travellers who screened at given time since infection. Columns show three possible mean incubation periods, and rows show three plausible probabilities that an infected person is subclinical. Here, we assume screening occurs at both arrival and departure; see Fig. 2 - supplementary figure 1 and Fig. 2 - supplementary figure 2 for departure or arrival screening only. The black dashed lines separate detected cases (below) from missed cases (above). Here, we assume flight duration = 24 hours, the probability that an individual is aware of exposure risk is 0.2, the sensitivity of fever scanners is 0.7, and the probability that an individual will truthfully self-report on risk questionnaires is 0.25. Table 1 lists all other input values.

Second, we considered a population of infected travellers, each with a unique time of exposure, and in turn a unique probability of having progressed to the symptomatic stage. Here, the model used a resampling-based approach to simultaneously consider uncertainty from both (1) stochasticity in any single individual’s screening outcome, and (2) uncertainty as to the true, underlying natural history parameters driving the epidemic. Details are provided in the methods, but briefly, we constructed 1000 plausible parameter sets, drawn using Latin hypercube sampling from plausible ranges for each parameter (Table 1). Using each parameter set, we simulated screening outcomes for a population of 100 infected individuals. Fig. 3A shows the distribution of infected travellers detected per simulation, and Fig. 3B shows the mean fraction of individuals with each screening outcome from across all simulations.

**Fig 3.**
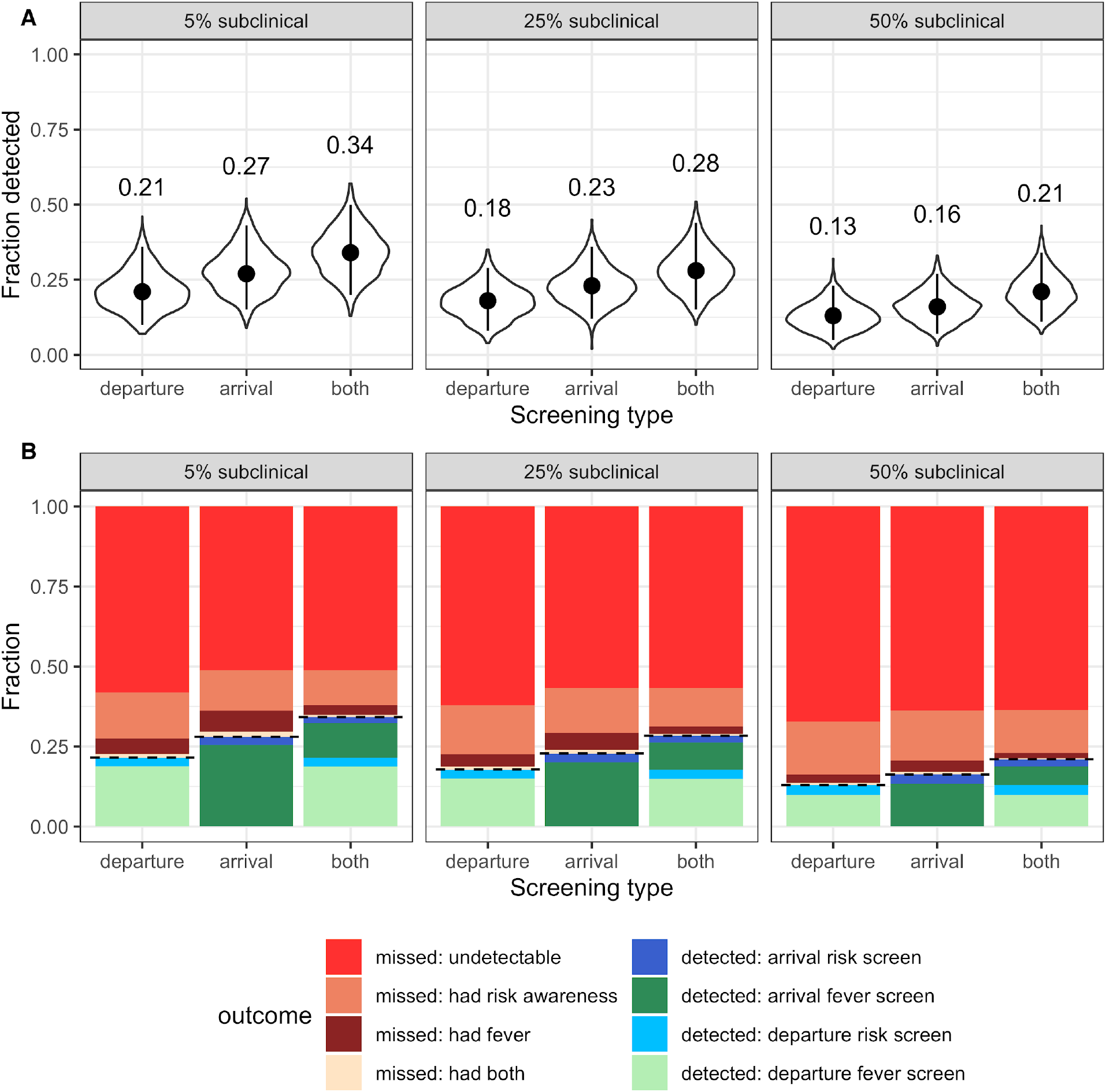
Population-level outcomes of screening programs in a growing epidemic. (A) Violin plots of the fraction of infected travellers detected, accounting for current uncertainties by running 1000 simulations using parameter sets randomly drawn from the ranges shown in Table 1. Dots and vertical line segments show the median and central 95%, respectively. Text above each violin shows the median fraction detected. (B) Mean fraction of travellers with each screening outcome. The black dashed lines separate detected cases (below) from missed cases (above).

### Individual probabilities of a given screening outcome

Our model outputs the probability of different screening outcomes through time, including the overall likelihood of detecting the infected traveller and the different contributions to success or failure. First, we explored the probability that any particular infected individual would be detected by a screening program, as a function of the time between exposure and the initiation of travel (Fig. 2). A crucial driver of the effectiveness of traveller screening programs is the duration of the incubation period, particularly since infected people are most likely to travel before the onset of symptoms. Here we considered three scenarios with different mean incubation periods: 5.5 days is most consistent with most existing estimates, while 4.5 and 6.5 days provide a sensitivity analysis roughly consistent with ranges, confidence or credible intervals reported elsewhere (Backer et al., 2020; Chan et al., 2020; Li et al., 2020; Liu et al., 2020). Even within the narrow range tested, screening outcomes were sensitive to the incubation period mean. For longer incubation periods, we found that larger proportions of departing travellers would not yet be exhibiting symptoms – either at departure or arrival – which in turn reduced the probability that screening would detect these cases, especially since we assume few infected travellers will realize they have been exposed to nCov.

A second crucial uncertainty is the proportion of cases that will develop detectable symptoms. We considered scenarios in which 5%, 25% and 50% of cases are subclinical, representing a best, middle and worst-case scenario, respectively. The middle and worst-case scenarios have predictable and discouraging consequences for the effectiveness of traveller screening, since they render large fractions of the population undetectable by fever screening (Fig. 2). Furthermore, mild cases who are unaware of their exposure risk are never detectable, by any means. This is manifested as the bright red ‘undetectable’ region which persists well beyond the mean incubation period. For a screening program combining departure and arrival screening, as shown in Fig. 2, the greatest contributor to case detection is the departure fever screen. The arrival fever screen is the next greatest contributor, with its value arising from two factors: the potential to detect cases whose symptom onset occurred during travel, and the potential to catch cases missed due to imperfect instrument sensitivity in non-contact infrared thermal scanners used in traveller screening (Table 1). Considering the effectiveness of departure or arrival screening only (Fig 2 - Supplementary figure 1-2), we see that fever screening is the dominant contributor in each case, but that the risk of missing infected travellers due to undetected fever is substantially higher when there is no redundancy from two successive screenings.

### Overall screening effectiveness in a population of infected travellers during a growing epidemic

Next we computed population-level estimates of the effectiveness of different screening programs, as well as the uncertainties arising from the current partial state of knowledge about this recently-emerged virus. To do so, we modeled plausible population-level outcomes by tracking the fraction of infected travellers detained, given a growing epidemic and current uncertainty around parameter values. We separately consider the best, middle and worst-case scenarios for the proportion of infections that are subclinical, and for each scenario we compare the impact of departure screening only, arrival screening only, or programs that include both.

The striking finding is that even under the best-case assumptions, with just one infection in twenty being subclinical and all travellers passing through departure and arrival screening, the median fraction of infected travellers detected is only 0.34, with 95% interval extending from 0.20 up to 0.50 (Fig. 3A). The total fraction detected is lower for programs with only one layer of screening, with arrival screening preferable to departure screening owing to the possibility of symptom onset during travel. Considering higher proportions of subclinical cases, the overall effectiveness of screening programs is further degraded, with a median of just one in ten infected travellers detected by departure screening in the worst-case scenario. The key driver of these poor outcomes is that, even in the best-case scenario, nearly two thirds of infected travellers will not be detectable (as shown by the red regions in Fig. 3B). This is because in a growing epidemic, the majority of travellers will have been recently infected and hence will not yet have progressed to the symptomatic stage, and because we assume that few are aware of their exposure risk. As above, the dominant contributor to successful detections is fever screening.

### Interactive online app for public use

We have developed an interactive web application using Shiny in which users can replicate our analyses using parameter inputs that reflect the most up-do-date information. The supplementary user interface can be accessed at https://faculty.eeb.ucla.edu/lloydsmith/screeningmodel. Please note that while the results in Fig. 3 consider a range of plausible values for each parameter, the outputs of the Shiny app are calculated using fixed, user-specified values only.

## Discussion

The international expansion of nCoV cases has led to travel screening measures being proposed and implemented in numerous countries. Given the rapid growth of the epidemic in China, emphasis on these measures is likely to rise in an attempt to prevent community spread of the virus in new geographic areas. Using a mathematical model of screening with preliminary estimates of nCoV epidemiology and natural history, we found that screening will in the best case only detect less than half of infected travellers. We found that two main factors influenced the effectiveness of screening. First, symptom screening depends on the natural history of an infection: individuals are increasingly likely to show detectable symptoms with increasing time since exposure. A fundamental challenge of screening is that many infected individuals will travel during their incubation period, a point at which they still feel healthy enough to travel but are simultaneously most difficult to detect. This effect is amplified when the incubation period is longer; infected individuals have a longer window in which they may travel with low probability of detection. Second, screening depends on whether exposure risk factors exist that would facilitate specific and reasonably sensitive case detection by questionnaire. For nCoV, there is so far limited evidence for specific risk factors; we therefore assumed that at most 40% of travellers would be aware of a potential exposure, and that a minority would self-report their exposure honestly, which led to limited effectiveness in questionnaire-based screening. The confluence of these two factors led to many infected travellers being fundamentally undetectable. Even under our most generous assumptions about the natural history of nCoV, the presence of undetectable travellers made the greatest contribution to screening failure. Correctable failures, such as missing a traveller with fever or awareness of their exposure risk, played a more minor role.

There are some limitations to our analysis. Parameter values for nCoV, such as the incubation period, are based on the limited data currently available. For such parameters, the tail of the distribution is important for understanding the potential for long delays until symptoms, but the tails of skewed distributions are notoriously difficult to characterize using limited data. In general, current parameter estimates may also be affected by bias or censoring, particularly in the early stages of an outbreak when most cases have been recently infected, and when data are primarily available for relatively severe, hospitalized cases. Another crucial uncertainty highlighted by our analysis is the frequency of cases too mild or non-specific to be detected as nCoV infections. At least one asymptomatic case is known to have occurred in a child (Chan et al., 2020). Further, children and young adults have been conspicuously underrepresented among hospitalized cases (Chen et al., 2020; Huang et al., 2020; Li et al., 2020). The possibility cannot be ruled out that large numbers of subclinical cases are occurring, especially in young people. If an age-by-severity interaction does indeed exist, then the mean age of travellers should be taken into account when estimating screening effectiveness. Further, transmission occurred before the onset of symptoms in one recent case report (Rothe et al., 2020). While it is too early to draw conclusions from a single case report, determining whether pre-symptomatic transmission is the norm also has major implications for the risk of establishing on-going spread in new locales.

As country-specific screening policies can change rapidly in real-time, we focused on a general screening framework rather than specific case studies. We also assumed traveller adherence and no active evasion of screening. However, there are informal reports of people taking antipyretics to beat fever screening (Mahbubani, 2020), which would further reduce the effectiveness of these methods. With travel restrictions in place, individuals may also take alternative routes (e.g. road rather than air), which would in effect circumvent departure and/or arrival screening as a control measure. Our quantitative findings may overestimate screening effectiveness if many travellers evade screening.

Our results have several implications for the design and implementation of control measures. Arrival screening could delay the introduction of cases if the infection is not yet present (Cowling et al., 2010), or reduce the initial rate of spread by limiting the number of parallel chains of transmission initially present in a country. But because screening is inherently leaky, it is crucial to also have measures in place to identify cases missed at arrival screening. For example, travellers could be provided with an information card to self-screen and self-report (Public Health England, n.d.), alongside increased general surveillance/alertness in healthcare settings. We should not take false confidence from reports that infected travellers are being detected by existing screening programs. Our findings indicate that for every case detected by travel screening, one or more infected travellers were not caught, and must be found and isolated by other means.

The expected high miss rate of screening programs also has implications for assessing when different programs are worthwhile investments. For areas yet to experience community-based transmission of the virus, and subject to substantial traveller inflows from affected areas, arrival screening can delay importation of cases and build awareness among incoming travellers. Even once there is some early-stage community transmission in a specific location, arrival screening may still reduce the chance of multiple independent transmission chains and ease the work of contact tracing teams, although the relative benefit of such screening for overall case prevention with decline as local transmission increases. Once there is generalized spread which has outpaced contact tracing, departure screening to prevent export of cases to new areas will be more valuable than arrival screening to identify additional incoming cases. However the cost-benefit tradeoff for any screening policy should be assessed in light of past experiences, where few or no infected travellers have been detected by such programs (Gostic et al., 2015).

Several factors could potentially strengthen the screening measures described here. With improved efficiency of thermal scanners or other symptom detection technology, we would expect a smaller difference between the effectiveness of arrival-only screening and combined departure and arrival screening in our analysis. Alternatively, the benefits of redundant screening (noted above for programs with departure and arrival screens) could be gained in a single-site screening program by simply having two successive fever-screening stations that travellers pass through (or taking multiple measurements of each traveller at a single station). As risk factors become better known, questionnaires could be refined to identify more potential cases. Alternatively, less stringent definition of high exposure risk (e.g. contact with anyone with respiratory symptoms) would be more sensitive. These approaches would boost sensitivity of screening, but could also incur a large cost in terms of false positives detained, especially during influenza season.

The availability of rapid PCR tests would also be beneficial for case identification at arrival, and would address concerns with false-positive detections. If such tests were fast, there may be potential to test suspected cases in real time based on questionnaire responses, travel origin, or borderline symptoms. However, such measures could prove highly expensive if implemented at scale. There is also scope for new tools to improve the ongoing tracking of travellers who pass through screening, such as smartphone-based self-reporting of temperature or symptoms in incoming cases. Recent travellers could even be asked to maintain a diary of close contacts for 14 days following arrival, to expedite contact tracing in the event they become ill with nCoV. This would be cheaper and more scalable than intense follow-up, but is likely to be limited by user adherence.

Our analysis underscores the reality that respiratory viruses are difficult to detect by travel screening programs, particularly if a substantial fraction of infected people show mild or indistinct symptoms, and if incubation periods are long. Quantitative estimates of screening effectiveness will improve as more is learned about this recently-emerged virus, and will vary with the precise design of screening programs. However, we present a robust qualitative finding: in any situation where there is widespread epidemic transmission in source populations from which travellers are drawn, travel screening programs can slow but not stop the importation of infected cases. By decomposing the factors leading to success or failure of screening efforts, our work supports decision-making about program design, and highlights key questions for further research. We hope that these insights may help to mitigate the global impacts of nCoV by guiding effective decision-making in both high- and low-resource countries, and may contribute to prospective improvements in travel screening policy for future emerging infections.

## Materials and Methods

### Modeling strategy

The model’s structure is summarized above (Fig. 1), and detailed methods have been described previously (Gostic et al., 2015). Here, we summarize relevant extensions, assumptions and parameter inputs.

### Extensions

Our previous model tracked all the ways in which infected travellers can be detected by screening (fever screen, or risk factor screen at arrival or departure). Here, we additionally keep track of the many ways in which infected travellers can be missed (i.e. missed given fever present, missed given exposure risk present, missed given both present, or missed given undetectable). Cases who have not yet passed the incubation period are considered undetectable by fever screening, even if they will eventually develop symptoms in the future. In other words, no traveller is considered “missed given fever present” until they have passed the incubation period and show detectable symptoms. Infected travellers who progress to symptoms during their journey are considered undetectable by departure screening, but detectable by arrival screening. Additionally, in the supplementary user interface, we implemented the possibility that some fraction of infected travellers deliberately evade screening.

### Fraction of subclinical cases

Our best-case scenario, in which only 5% of cases are subclinical, is consistent with the fact that the vast majority of nCoV cases detected to date have shown fever or other detectable symptoms (Chan et al., 2020; Chen et al., 2020; Huang et al., 2020). But so far the data have primarily captured severe, hospitalized cases, so the true fraction of subclinical nCoV cases remains a crucial unknown. Particularly given the conspicuous under-representation of children and young adults among hospitalized patients (Huang et al., 2020; Li et al., 2020), our medium and worst-case scenarios (75% and 50% subclinical) remain plausible.

### Incubation period distribution

Numerous recent studies have estimated that the incubation period lasts about 5.5 days on average (Backer et al., 2020; Chan et al., 2020; Li et al., 2020; Liu et al., 2020), with the tail of the distribution stretching to at least 12 days (Backer et al., 2020; Li et al., 2020). Consistent with these observations, a recent study by Backer, Klinkberg and Wallinga (2020) characterized the incubation period distribution for nCoV, concluding that a Weibull distribution provided the best fit to data, but that a gamma distribution performed almost as well. We proceed by adopting their best-fit gamma distribution (mean 5.7 days, s.d. 2.6, or alternatively, shape = 4.8, scale = 1.2), as the gamma form is more computationally convenient within our model. In order to vary the mean incubation period in our uncertainty analyses while maintaining the shape of this two-parameter distribution, we fix the scale parameter to 1.2, and set the shape parameter equal to 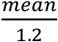 (Fig. 3 - supplementary figure 1).

### Effectiveness of exposure risk questionnaires

The probability that an infected traveller is detectable using questionnaire-based screening for exposure risk will be highest if specific risk factors are known. Other than close contact with a known nCoV case, or contact with the Hunan seafood wholesale market in the earlier phase of the outbreak in Wuhan, we are not aware that any specific risk factors have been identified. Given the relative anonymity of respiratory transmission, we assume that a minority of infected travellers would realize that they have been exposed before symptoms develop (20% in Fig. 2, range 5-40% in Fig. 3). Further, relying on a previous upper-bound estimate (Gostic et al., 2015) we assume that only 25% of travellers would self-report truthfully if aware of elevated exposure risk.

Table 1 summarizes the state of knowledge about additional key natural history parameters, as of January 28, 2020.

## Data Availability

All relevant code is available at https://github.com/kgostic/traveller_screening.
The model does not input any case data.
All relevant inputs are present in the code, and described in Table 1 or in the manuscript text.

https://faculty.eeb.ucla.edu/lloydsmith/screeningmodel

## Acknowledgements

We thank the Cobey lab for helpful comments.

## Code and data availability

All code used in these analyses can be found at https://github.com/kgostic/traveller_screening.

## Supplementary Figures

**Fig 2-Supplementary figure 1.**
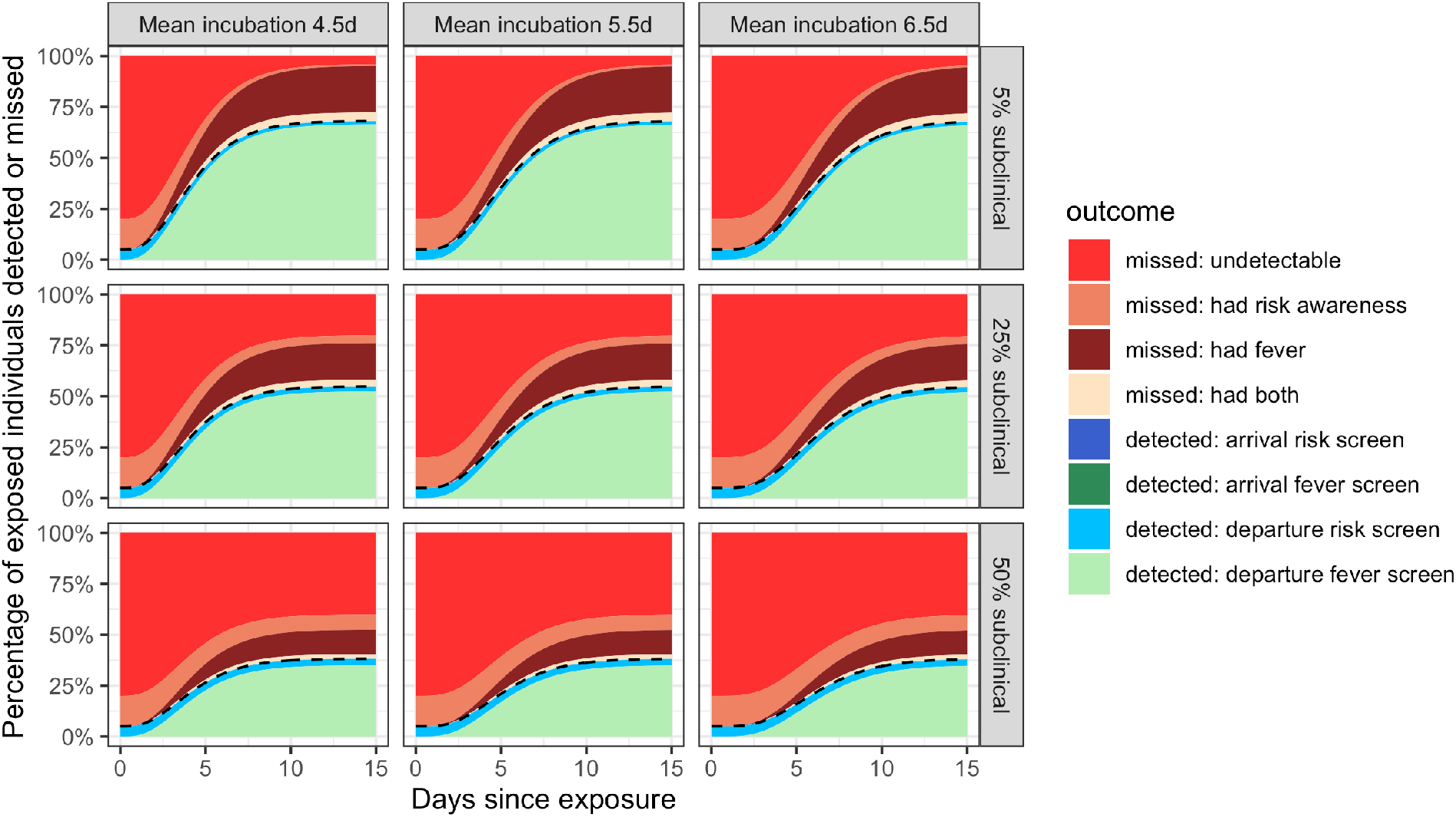
Departure screening only.

**Fig 2-Supplementary figure 2.**
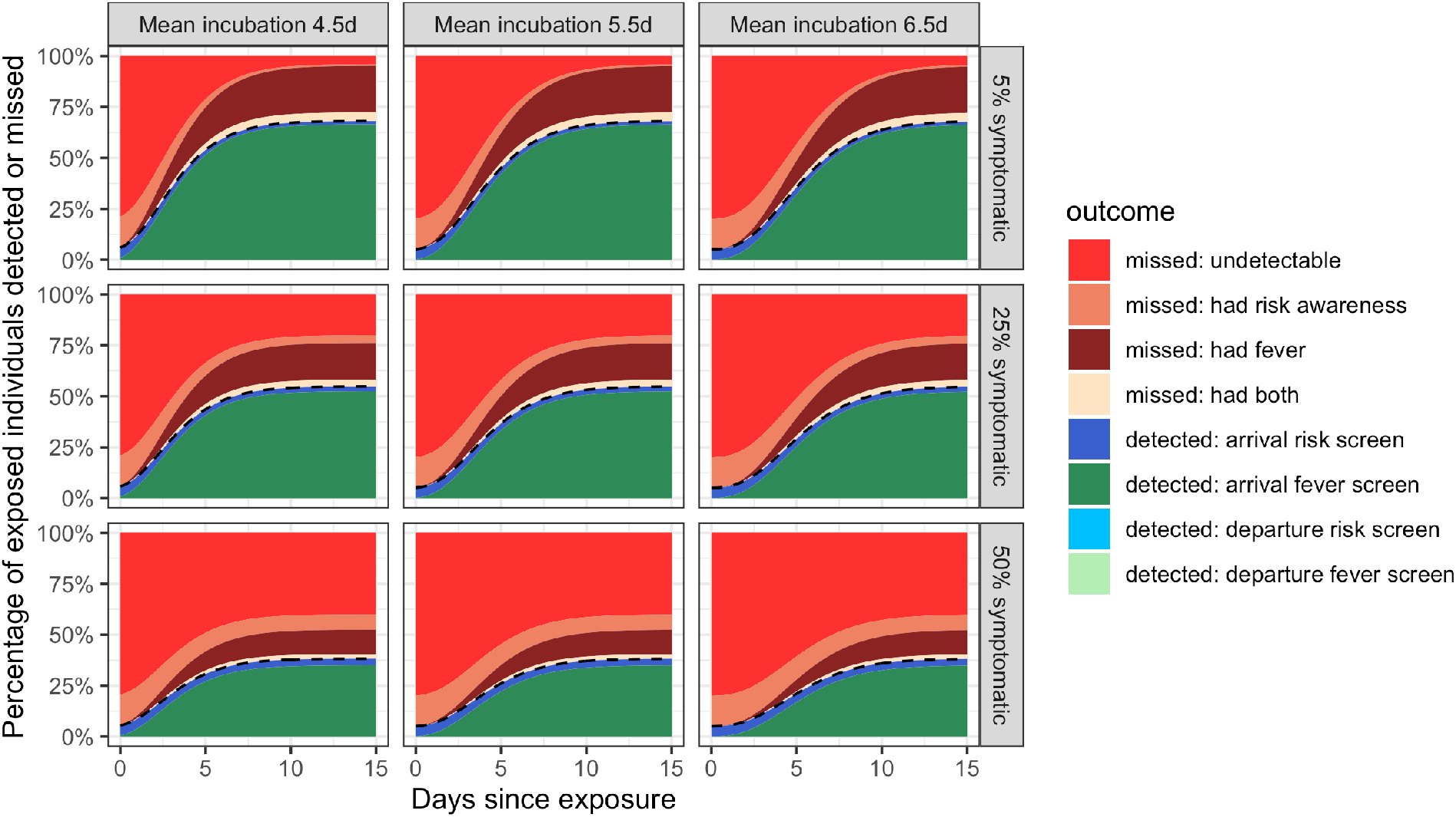
Arrival screening only.

**Fig 3- Supplementary figure 1.**
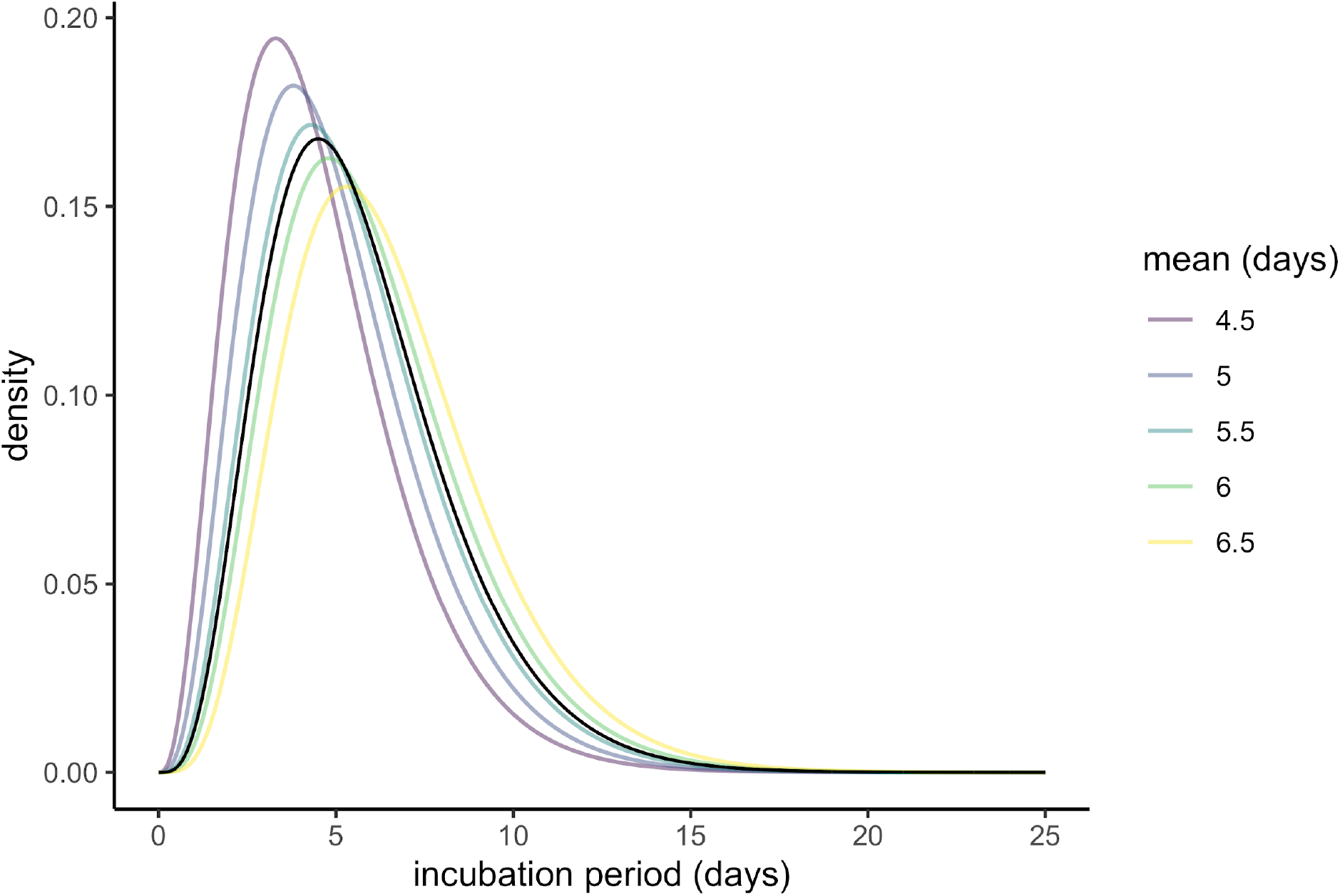
Plausible incubation period distributions underlying the analyses in Fig. 3. The black line shows the probability density function of the best-fit gamma distribution reported by (Backer et al., 2020). Other lines show the probability density functions for different assumptions regarding the mean incubation period. Each is a gamma distribution with scale = 1.2, and shape = 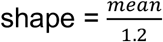.

## References

Backer JA, Klinkenberg D, Wallinga J. 2020. The incubation period of 2019-nCoV infections among travellers from Wuhan, China. medRxiv 2020.01.27.20018986. doi:10.1101/2020.01.27.20018986

Bitar D, Goubar A, Desenclos JC. 2009. International travels and fever screening during epidemics: A literature review on the effectiveness and potential use of non-contact infrared thermometers. Eurosurveillance 14:1–5.

Chan JF, Yuan S, Kok K, To KK, Chu H, Yang J, Xing F, Liu J, Yip CC, Poon RW, Tsoi H, Lo SK, Chan K, Poon VK, Chan W, Ip JD, Cai J-P, Cheng VC-C, Chen H, Hui CK-M, Yuen K-Y. 2020. Articles A familial cluster of pneumonia associated with the 2019 novel coronavirus indicating person-to-person transmission?: a study of a family cluster. The Lancet 1–10. doi:10.1016/S0140-6736(20)30154-9

Chen N, Zhou M, Dong X, Qu J, Gong F, Han Y, Qiu Y, Wang J, Liu Y, Wei Y, Xia J, Yu T, Zhang X, Zhang L. 2020. Epidemiological and clinical characteristics of 99 cases of 2019 novel coronavirus pneumonia in Wuhan, China: a descriptive study. The Lancet 0. doi:10.1016/S0140-6736(20)30211-7

Cowling BJ, Lau LL, Wu P, Wong HW, Fang VJ, Riley S, Nishiura H. 2010. Entry screening to delay local transmission of 2009 pandemic influenza A (H1N1). BMC Infect Dis 10:82. doi:10.1186/1471-2334-10-82

Gostic KM, Kucharski AJ, Lloyd-Smith JO. 2015. Effectiveness of traveller screening for emerging pathogens is shaped by epidemiology and natural history of infection. eLife 2015:1–16. doi:10.7554/eLife.05564

Huang C, Wang Y, Li X, Ren L, Zhao J, Hu Y, Zhang L, Fan G, Xu J, Gu X, Cheng Z, Yu T, Xia J, Wei Y, Wu W, Xie X, Yin W, Li H, Liu M, Xiao Y, Gao H, Guo L, Xie J, Wang G, Jiang R, Gao Z, Jin Q, Wang J, Cao B. 2020. Articles Clinical features of patients infected with 2019 novel coronavirus in Wuhan, China. The Lancet 1–10. doi:10.1016/S0140-6736(20)30183-5

Imai N, Cori A, Dorigatti I, Baguelin M, Donnelly CA, Riley S, Neil M. 2019. Report 3: Transmissibility of 2019-nCoV. London.

Kucharski AJ, Russell T, Diamond C, Funk S, Eggo R. n.d. Analysis of early transmission of 2019-nCoV and implications for outbreaks in new locations.

Li Q, Guan X, Wu P, Wang X, Zhou L, Tong Y, Ren R, Leung KSM, Lau EHY, Wong JY, Xing X, Xiang N, Wu Y, Li C, Chen Q, Li D, Liu T, Zhao J, Liu M, Tu W, Chen C, Jin L, Yang R, Wang Q, Zhou S, Wang R, Liu H, Luo Y, Liu Y, Shao G, Li H, Tao Z, Yang Y, Deng Z, Liu B, Ma Z, Zhang Y, Shi G, Lam TTY, Wu JT, Gao GF, Cowling BJ, Yang B, Leung GM, Feng Z. 2020. Early Transmission Dynamics in Wuhan, China, of Novel Coronavirus–Infected Pneumonia. N Engl J Med 0:null. doi:10.1056/NEJMoa2001316

Liu T, Hu J, Kang M, Lin L, Zhong H, Xiao J, He G, Song T, Huang Q, Rong Z, Deng A, Zeng W, Tan X, Zeng S, Zhu Z, Li J, Wan D, Lu J, Deng H, He J, Ma W. 2020. Transmission dynamics of 2019 novel coronavirus (2019-nCoV). bioRxiv 1–13.

Priest PC, Duncan AR, Jennings LC, Baker MG. 2011. Thermal Image Scanning for Influenza Border Screening: Results of an Airport Screening Study. PLOS ONE 6:e14490. doi:10.1371/journal.pone.0014490

Read JM, Bridgen JRE, Cummings DAT, Ho A, Jewell CP. 2020. Novel coronavirus 2019-nCoV: early estimation of epidemiological parameters and epidemic predictions. medRxiv. doi:10.1101/2020.01.23.20018549

Riou J, Althaus CL. 2020. Pattern of early human-to-human transmission of Wuhan 2019-nCoV. bioRxiv 1–6.

Tay MR, Low YL, Zhao X, Cook AR, Lee VJ. 2015. Comparison of Infrared Thermal Detection Systems for mass fever screening in a tropical healthcare setting. Public Health 129:1471–1478. doi:10.1016/j.puhe.2015.07.023

World Health Organization. n.d. Novel coronavirus (2019-nCov) Situation Report 12. https://www.who.int/docs/default-source/coronaviruse/situation-reports/20200201-sitrep-12-ncov.pdf?sfvrsn=273c5d35_2

Wu JT, Leung K, Leung GM. 2020. Nowcasting and forecasting the potential domestic and international spread of the 2019-nCoV outbreak originating in Wuhan, China: a modelling study. The Lancet 0. doi:10.1016/S0140-6736(20)30260-9

